# COVID-19 among adults living with HIV: Correlates of mortality in a general population in a resource-limited setting

**DOI:** 10.1101/2022.10.17.22281085

**Authors:** Reshma Kassanjee, Mary-Ann Davies, Olina Ngwenya, Richard Osei-Yeboah, Theuns Jacobs, Erna Morden, Venessa Timmerman, Stefan Britz, Marc Mendelson, Jantjie Taljaard, Julien Riou, Andrew Boulle, Nicki Tiffin, Nesbert Zinyakatira

**Affiliations:** School of Public Health, University of Cape Town, Cape Town, South Africa; Western Cape Government: Health and Wellness, Cape Town, South Africa; Wellcome Centre for Infectious Disease Research in Africa, Institute of Infectious Diseases and Molecular Medicine, University of Cape Town, Cape Town, South Africa; Division of Computational Biology, Integrative Biomedical Sciences Department, Faculty of Health Sciences, University of Cape Town, Cape Town, South Africa; Department of Statistical Sciences, University of Cape Town, Cape Town, South Africa; Division of Infectious Diseases and HIV Medicine, Department of Medicine, Groote Schuur Hospital, University of Cape Town, South Africa; Division of Infectious Diseases, Department of Medicine, Tygerberg Hospital, Stellenbosch University, South Africa; Institute of Social and Preventive Medicine, University of Bern, Bern, Switzerland; University of the Western Cape, Cape Town, South Africa

**Keywords:** HIV, SARS-CoV-2, COVID-19, mortality, CD4 count, viral load, South Africa

## Abstract

**Introduction:** While a large proportion of people with HIV (PWH) have experienced SARS-CoV-2 infections, there is uncertainty about the role of HIV disease severity on COVID-19 outcomes, especially in lower income settings. We studied the association between mortality and characteristics of HIV severity and management, and vaccination, among adult PWH.

**Methods:** We analysed observational cohort data on all PWH aged ≥15 years experiencing a diagnosed SARS-CoV-2 infection (until March 2022), who accessed public sector healthcare in the Western Cape province of South Africa. Logistic regression was used to study the association of mortality with CD4 cell count, viral load, evidence of ART, time since first HIV evidence, and vaccination, adjusting for demographic characteristics, comorbidities, admission pressure, location and time period.

**Results:** Mortality occurred in 5.7% (95% CI: 5.3,6.0) of 17 831 first diagnosed infections. Higher mortality was associated with lower recent CD4, no evidence of ART collection, high or unknown recent viral load (among those with ART evidence), and recent first HIV evidence, differentially by age. Vaccination was protective. The burden of comorbidities was high, and tuberculosis, chronic kidney disease, diabetes and hypertension were associated with higher mortality, more strongly in younger adults.

**Conclusions:** Mortality was strongly associated with suboptimal HIV control, and prevalence of these risk factors increased in later COVID-19 waves. It remains a public health priority to ensure PWH are on suppressive ART and vaccinated, and manage any disruptions in care that occurred during the pandemic. The diagnosis and management of comorbidities, including for tuberculosis, should be optimised.

## Introduction

More than two years have passed since the start of the COVID-19 pandemic. A large proportion of people with HIV (PWH) have experienced SARS-CoV-2 infections, and this population appears to havea modestly increased risk of severe COVID-19 outcomes [1-7]. However, sincemost published research has been from high income settings with almost all PWH on virally suppressive antiretroviral therapy (ART), there is still uncertainty about the role of HIV disease severity on COVID-19 outcomes and the extent of mitigation with effective ART, especially in lower income settings [1]. The recent WHO Global Clinical Platform of COVID-19 study showed that being on ART and/or virally suppressed was protective against poor COVID-19 outcomes in hospitalized patients, but there is limited data on all COVID-19 patients including those not admitted, and the effect of immunosuppression before the COVID-19 episode [7, 8]. Furthermore, the effectiveness of vaccination in preventing severe COVID-19 in PWH in this context remains unclear [9, 10].

A better understanding of the risk factors for severe COVID-19 in PWH is vital to guide optimal use of anti-SARS-CoV-2 therapies, especially in resource-limited settings where access to these therapies remains extremely limited, but the majority of immunosuppressed and/or viraemic PWH live [11, 12]. Intermittent surges of COVID-19 infections are likely to continue as the virus mutates, especially since most countries have relaxed COVID-19 restrictions due to their unsustainable socio-economic cost or most people now being protected against severe disease by vaccination and/or prior infection. Severe COVID-19 may also be more prolonged in PWH, facilitating accumulation of mutations with a risk of variant evolution [13, 14]

We aimed to study the association between mortality and characteristics of HIV severity (CD4 cell count and HIV viral load (VL)) and management (ART collection and time since first HIV evidence) as well as SARS-CoV-2 vaccination status among adult (aged ≥15 years) PWH who experienced a diagnosed SARS-CoV-2 infection in the Western Cape (WC) province of South Africa (SA), adjusted for demographic characteristics, comorbidities, admission pressure, location and time period.

## Methods

### Study Population and Data Sources

We studied adults (aged ≥15 years) with both HIV and SARS-CoV-2 infections who utilised public sector healthcare in the WC. The WC has about 5.5 million people aged ≥15 years with estimated HIV prevalence of 14.2% among women and 7.7% among men [15, 16]. Approximately 75% of the population are dependent on public sector health services, but a greater proportion amongst PWH [17].

De-identified data was extracted from the Western Cape Provincial Health Data Centre (WCPHDC) which uses a unique patient identifier to integrate patient-level data from multiple information systems (administrative, laboratory, pharmacy, and disease management) in all public sector health facilities in the province [17]. We included all adults who had accessed healthcare in the three years preceding the start of the COVID-19 epidemic in SA (1 March 2020), a laboratory-confirmed positive SARS-CoV-2 PCR or antigen test in the public or private sector, and evidence of HIV before or less than 3 weeks after the SARS-CoV-2 diagnosis. We included all SARS-CoV-2 diagnoses until 10 March 2022 and data was extracted on 28 April 2022 to allow for sufficient time to ascertain mortality. HIV and other comorbidities were inferred from a range of sources including laboratory tests, electronic disease management systems for HIV and tuberculosis, pharmacy data dispensed for specific conditions (e.g., combination ART for HIV and anti-diabetic medication for diabetes) and ICD-10 diagnostic codes. Each inferred comorbidity was assigned a ‘first evidence date’ which was the earliest date on which any source indicated that the condition was present. The WCPHDC links data on people with diagnosed SARS-CoV-2 infection to the vital registration system on deaths weekly, using national identification numbers, and reviews all COVID-19 related deaths daily.

We obtained vaccination data by linking the South African national identifier to the Electronic Vaccine Data System which records all SARS-CoV-2 vaccines administered nationally. The following vaccines were available as a primary series to the general population from 17 May 2021, starting with those aged ≥60 years, with progressive access for younger age groups: Janssen/Johnson & Johnson (Ad26.COV2.S) (initially as a single dose for those aged ≥18 years, with second doses available from January 2022) and two doses of Pfizer-BioNTech (BNT162b2). By 20 October 2021, everyone aged ≥12 years was eligible for vaccination. Immunocompromised individuals including PWH with CD4<200 cells/µl were eligible for an additional homologous vaccine dose (i.e. a second Ad26.COV2.S or third BNT162b2), while further booster doses were available to all aged ≥18 years 180 days after the previous dose from January 2022.

The study was approved by the University of Cape Town Health Research Ethics committee and the Western Cape Government Provincial Department of Health.

### Definitions of the mortality outcome, HIV characteristics, and covariates

An outcome of death was recorded if death occurred either within 14 days before to 28 days after the date of SARS-CoV-2 diagnosis or within 14 days after discharge from a hospitalization that started within 21 days of the SARS-CoV-2 diagnosis, and there was no clear non-COVID-19 cause of death recorded.

For the HIV health characteristics, for CD4 cell count, from amongst all measurements obtained earlier than two weeks before the SARS-CoV-2 diagnosis, we extracted the most recent CD4 within the previous 18 months, and, for secondary analyses, the CD4 nadir and CD4 at ART initiation (closest to, and within six months before and two weeks after, ART initiation). For ART, we considered whether there is any evidence of ART collection earlier than three days before SARS-CoV-2 diagnosis. For VL, among those with any ART evidence, we used the most recent measurement obtained within the 24 months before and two weeks after the SARS-CoV-2 diagnoses. Time since first HIV evidence was also considered.

Demographic covariates included were sex and age. We included indicators of the chronic conditions diabetes, hypertension, chronic kidney disease (CKD), and chronic obstructive pulmonary disease (COPD). For each of these, the first evidence date could occur up to three months after SARS-CoV-2 diagnosis, except for COPD where the first evidence needed to occur earlier than two weeks before the SARS-CoV-2 diagnosis (later evidence may be a consequence of COVID-19 or incorrectly inferred from medication prescribed during the COVID-19 episode). Time since the most *recent* evidence of the start of a tuberculosis episode (not more than one month after the SARS-CoV-2 diagnosis) was also considered, with episodes beginning after two months before the SARS-CoV-2 diagnosis considered as ongoing; as well as an indicator of current pregnancy.

Vaccination status at the time of SARS-CoV-2 diagnosis was assigned the highest level of protection from among six categories, based on time since and number of vaccines already received: No vaccine received, <28 days since a first dose of either vaccine type (‘early’ vaccination), ≥28 days since a first dose of a vaccine (either Janssen or Pfizer–BioNTech, separately), or ≥14 days since a second dose (by type). Below we sometimes combine categories indicating the primary series (two Pfizer doses or ≥1 Janssen dose). No persons had third vaccines during our analysis period; boosters were not widely available until later.

Variation by location and time were described by district, where the province is sub-divided into six districts, and wave period, where the four distinct waves coincided with periods of different variants being dominant in the WC, namely the ancestral, Beta, Delta and Omicron BA.1/BA.2 variants respectively (wave 1: 2 May - 6 Aug 2020; wave 2: 13 Nov 2020 - 1 Feb 2021; wave 3: 3 June - 19 Sep 2021; wave 4: 28 Nov 2021 - 24 Feb 2022; inter-wave periods: remaining time; where waves were defined as the periods when the 7-day moving average of new COVID-19 diagnoses was ≥ 60/million population). Additionally, we constructed a time-specific proxy of availability of healthcare: per district, admission pressure was the week’s total number of COVID-19 admissions relative to the district’s maximum weekly COVID-19 admissions (as a percentage).

### Statistical Analysis

Data management and analyses were conducted in SQL server Management Studio (version 8), STATA (version 15.1) and R (version 4.05).

In the primary analysis, we included infections only until the end of wave 3 because of concerns that the SARS-CoV-2 diagnosis may be co-incidental in a sizable proportion of deaths in those with SARS-CoV-2 infection in the fourth (Omicron BA.1/BA.2) wave when the prevalence of mild/asymptomatic COVID-19 was high and there was extensive testing of all hospitalized patients [18]. Because reinfection risk was minimal until the end of the (third) Delta wave [3], only first documented infections per person were included in the regression analyses described below, also to ensure statistical independence of data points.

We used multivariable logistic regression to assess the association between mortality and HIV characteristics (CD4 cell count, viral load, ART collection, time since first HIV evidence) as well as vaccination, adjusted for the person’s characteristics (sex, age, diabetes, hypertension, CKD, COPD, time since tuberculosis, and pregnancy) and time and location (district, wave period, and admission pressure). In the primary models, from among the three CD4 measures, we used the most recent CD4. We included all terms as main effects and used likelihood ratio tests to obtain covariate-level p-values. To improve model-fit (assessed by comparing observed to model-expected proportions experiencing death), we fitted separate models to each of two age groups(15-39 years versus≥ 40 years old) and included as an additional covariate an indicator of whether the total number of co-existing conditions (chronic conditions and ongoing/previous tuberculosis) was greater than two.

As supplementary content, in secondary analyses, alternative models used either the CD4 nadir or CD4 at ART initiation, instead of the most recent CD4; or analysed all (first) cases until 10 March 2022.

## Results

About 4.2% of the nearly 600 000 adults with HIV in the WC who accessed public health services in the 3 years before March 2020 were diagnosed with SARS-CoV-2 infection by 10 March 2022. While the general population experienced much higher infection peaks during subsequent COVID-19 waves compared to the first, waves 1 to 4 in adult PWH had more consistent peaks (Figure 1A), with a much steeper rise in infections during the first wave, as well as the most infections that occurred in PWH being registered during this wave (see SDC A for additional descriptive statistics). The risk of mortality differed by HIV characteristics and vaccination status (Figure 1B).

**Figure 1:**
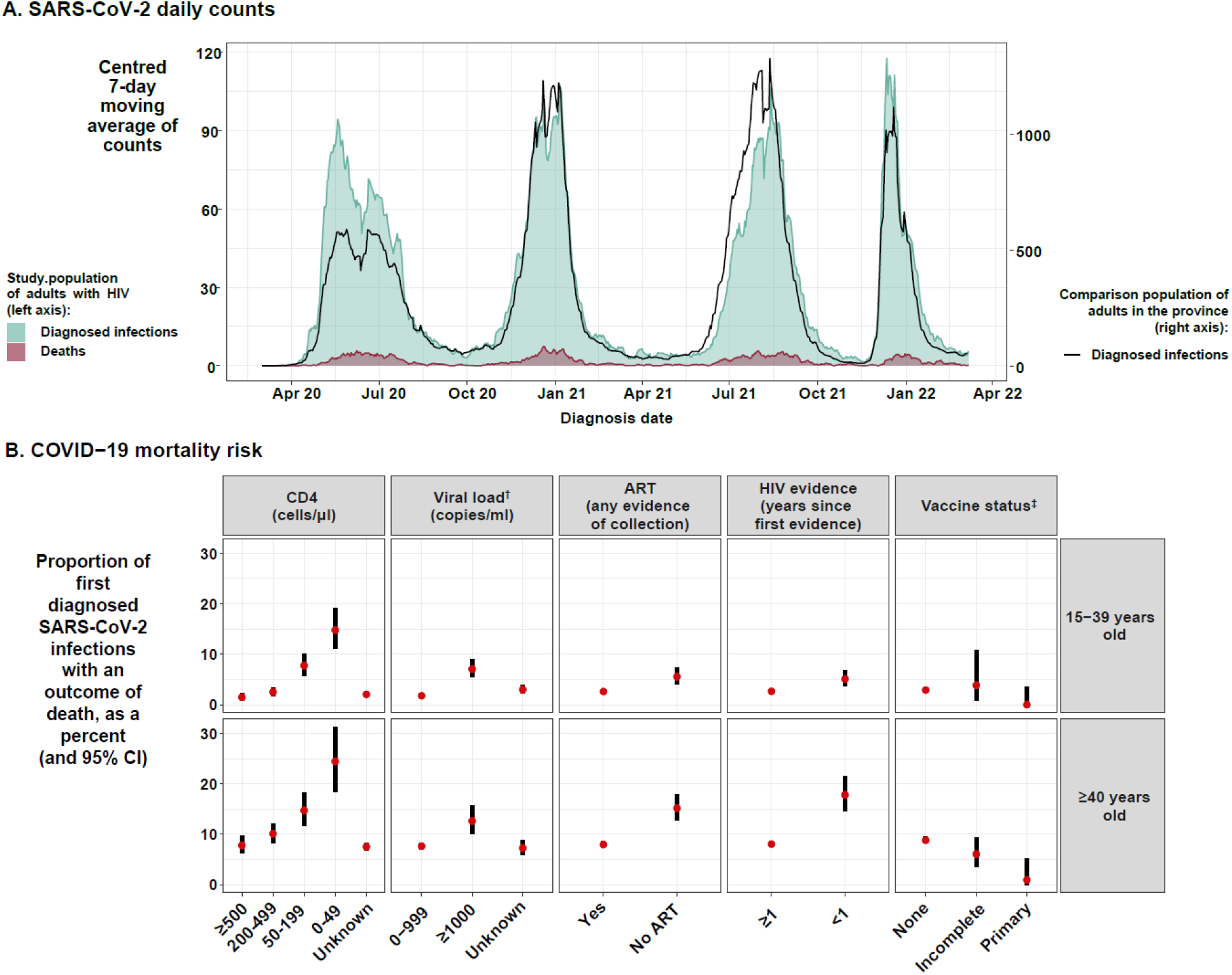
Description of SARS-CoV-2 infections and deaths among adults with HIV accessing public sector healthcare in the Western cape province: (A) counts of diagnosed SARS-CoV-2 infections and deaths over time, compared to diagnosed infections in the general population; and (B) the risk of mortality by HIV characteristics and vaccination in people who are experiencing a first diagnosed infection up to 10 March 2022, stratified by age. ^†^ Among those with evidence of ART. ^‡^ For descriptive purposes, ≥28 days since 1 Janssen vaccine, possibly with a 2^nd^ vaccine too, and ≥14 days since a 2nd Pfizer vaccine, were combined into the ‘primary’ series, and ‘early vaccination’ (<28 days since a vaccine) or 1 Pfizervaccine combined into ‘incomplete’. ART: Antiretroviral therapy. CI: Confidence interval.

We recorded 18 120 SARS-CoV-2 infections during the first three COVID-19 waves. Among these infections, 5.7% (1 027) were associated with death (Table 1). The median age of infected persons was 39 years (interquartile range IQR: 32,47) and nearly three quarters (71.0%) were female. Half (51.4%) of infections were in PWH with ≥1 other chronic conditions or ongoing/previous tuberculosis, and 15.5% of persons had multiple such conditions. Tuberculosis (ongoing/previous) was experienced by three in 10 (29.9%) persons, and hypertension was the most prevalent chronic condition (20.5%), followed by diabetes and COPD (9.7% and 7.2%). Chronic conditions were up to 7 times more prevalent in those aged ≥40 years versus younger.

**Table 1:** Outcomes, demographic characteristics and co-existing conditions for SARS-CoV-2 infections among adults with HIV in the Western Cape, stratified by age group (percentages and counts) ^†^ Time since the most recent evidence of a start of an episode; with episodes beginning after 2 months before the SARS-CoV-2 diagnosis considered as ongoing. ^‡^ Hypertension, diabetes, chronic obstructive pulmonary disease, chronic kidney disease, tuberculosis (at any time).

| Characteristic / Outcome | Cases until end of Wave 3 |  |  | All cases<br>(including Wave 4) |
| --- | --- | --- | --- | --- |
|  | Ages 15-39 years<br>n = 9,321 | Ages ≥40 years<br>n = 8,799 | All ages<br>n = 18,120 | All ages<br>n = 22,057 |
| **Death** | 2.9 (270) | 8.6 (757) | 5.7 (1,027) | 5.6 (1,233) |
| **Sex** |  |  |  |  |
| Female | 77.8 (7,249) | 63.8 (5,610) | 71.0 (12,859) | 70.5 (15,561) |
| Male | 22.2 (2,072) | 36.2 (3,189) | 29.0 (5,261) | 29.5 (6,496) |
| **Age (years)** |  |  |  |  |
| 15-24 | 9.6 (895) | - | 4.9 (895) | 5.0 (1,106) |
| 25-29 | 21.6 (2,011) | - | 11.1 (2,011) | 11.4 (2,517) |
| 30-34 | 32.8 (3,057) | - | 16.9 (3,057) | 17.4 (3,841) |
| 35-39 | 36.0 (3,358) | - | 18.5 (3,358) | 18.7 (4,130) |
| 40-49 | - | 60.1 (5,286) | 29.2 (5,286) | 28.6 (6,308) |
| 50-59 | - | 29.4 (2,590) | 14.3 (2,590) | 13.9 (3,057) |
| 60-69 | - | 8.2 (722) | 4.0 (722) | 3.9 (864) |
| ≥ 70 | - | 2.3 (201) | 1.1 (201) | 1.1 (234) |
| **Hypertension** | 8.7 (809) | 33.1 (2,914) | 20.5 (3,723) | 19.8 (4,360) |
| **Diabetes** | 4.0 (369) | 15.8 (1,391) | 9.7 (1,760) | 9.1 (1,999) |
| **Chronic obstructive pulmonary disease** | 4.1 (379) | 10.4 (917) | 7.2 (1,296) | 7.8 (1,714) |
| **Chronic kidney disease** | 0.9 (86) | 6.6 (584) | 3.7 (670) | 3.6 (784) |
| **Tuberculosis<sup>†</sup> (years since)** |  |  |  |  |
| Absent | 73.3 (6,830) | 66.7 (5,868) | 70.1 (12,698) | 68.3 (15,068) |
| ≥ 1 | 18.0 (1,674) | 26.4 (2,323) | 22.1 (3,997) | 22.3 (4,911) |
| < 1 | 3.6 (338) | 2.7 (238) | 3.2 (576) | 3.7 (806) |
| Ongoing | 5.1 (479) | 4.2 (370) | 4.7 (849) | 5.8 (1,272) |
| **Pregnancy (in women aged 15-49 years)** | 10.8 (784) | 1.4 (50) | 7.7 (834) | 7.8 (1,028) |
| **Number of previous SARS-CoV-2 diagnoses** |  |  |  |  |
| 0 | 98.1 (9,146) | 98.6 (8,675) | 98.3 (17,821) | 96.6 (21,316) |
| 1 | 1.9 (173) | 1.4 (121) | 1.6 (294) | 3.3 (722) |
| 2 | 0.0 (2) | 0.0 (3) | 0.0 (5) | 0.1 (19) |
| **Number of conditions<sup>‡</sup>** |  |  |  |  |
| 0 | 62.4 (5,814) | 36.0 (3,169) | 49.6 (8,983) | 48.7 (10,746) |
| 1 | 31.7 (2,957) | 38.3 (3,368) | 34.9 (6,325) | 35.7 (7,872) |
| 2 | 5.1 (476) | 17.9 (1,572) | 11.3 (2,048) | 11.5 (2,533) |
| ≥3 | 0.8 (74) | 7.8 (690) | 4.2 (764) | 4.1 (906) |

During the first three waves, very few of the people experiencing an infection (1.3%) had received their primary vaccination series (Table 2). For the HIV characteristics, a recent CD4 cell count was available for only 36.6% of persons, with median value of 446 (IQR: 274,648) cells/µL. Nearly one in 10 (9.1%) infected persons had never collected ART. Among those with any evidence of ART, in every 10 persons, one (9.1%) had a most recent VL ≥1000 copies/ml, while two (19.0%) had no recent measure. The median time since first HIV evidence was 6.3 years (IQR: 3.4,10.2).

**Table 2:** HIV characteristics and vaccination status for SARS-CoV-2 infections among adults with HIV in the Western Cape, stratified by age group (percentages and counts) ^†^ Among those with evidence of ART at any time. ^‡^ The highest level of protection from among six categories: No vaccine received (‘None’); <28 days since a first vaccine dose (‘Early’); ≥28 days since a first dose of a vaccine, by type (‘1 vaccine’); or ≥14 days since a second dose, by type (‘2 vaccines’). ART: Antiretroviral therapy

| Characteristic | Cases until end of Wave 3 |  |  | All cases<br>(including Wave 4) |
| --- | --- | --- | --- | --- |
|  | Ages 15-39 years<br>n = 9,321 | Ages ≥40 years<br>n = 8,799 | All ages<br>n = 18,120 | All ages<br>n = 22,057 |
| **CD4 count (cells/μl)** |  |  |  |  |
| ≥ 500 | 14.0 (1,304) | 11.3 (991) | 12.7 (2,295) | 12.6 (2,775) |
| 200-499 | 15.7 (1,465) | 11.5 (1,014) | 13.7 (2,479) | 13.5 (2,986) |
| 50-199 | 7.0 (648) | 5.8 (511) | 6.4 (1,159) | 7.1 (1,576) |
| 0-49 | 3.6 (333) | 2.2 (191) | 2.9 (524) | 3.5 (773) |
| Unknown | 59.8 (5,571) | 69.2 (6,092) | 64.4 (11,663) | 63.2 (13,947) |
| **Viral load† (copies/ml)** |  |  |  |  |
| 0-999 | 66.4 (5,642) | 77.7 (6,196) | 71.9 (11,838) | 69.6 (13,913) |
| ≥ 1000 | 10.8 (915) | 7.3 (585) | 9.1 (1,500) | 10.0 (2,003) |
| Unknown | 22.8 (1,941) | 15.0 (1,194) | 19.0 (3,135) | 20.4 (4,068) |
| **ART (years since most recent collection)** |  |  |  |  |
| ≥ 2 | 44.2 (4,116) | 46.5 (4,092) | 45.3 (8,208) | 46.0 (10,146) |
| < 2 | 47.0 (4,382) | 44.1 (3,883) | 45.6 (8,265) | 44.6 (9,838) |
| No ART | 8.8 (823) | 9.4 (824) | 9.1 (1,647) | 9.4 (2,073) |
| **HIV evidence (years since first)** |  |  |  |  |
| <1 | 9.0 (838) | 5.9 (519) | 7.5 (1,357) | 7.7 (1,706) |
| 1-5 | 39.2 (3,653) | 22.8 (2,002) | 31.2 (5,655) | 30.9 (6,826) |
| 5-10 | 36.5 (3,404) | 33.9 (2,986) | 35.3 (6,390) | 35.3 (7,781) |
| ≥ 10 | 15.3 (1,426) | 37.4 (3,292) | 26.0 (4,718) | 26.0 (5,744) |
| **Vaccination status‡** |  |  |  |  |
| None | 97.9 (9,126) | 95.2 (8,378) | 96.6 (17,504) | 89.6 (19,761) |
| Early | 0.8 (74) | 2.6 (126) | 1.6 (300) | 1.7 (379) |
| Janssen - 1 vaccine | 1.2 (111) | 1.1 (98) | 1.2 (209) | 3.7 (819) |
| Pfizer - 1 vaccine | 0.1 (9) | 0.9 (82) | 0.5 (91) | 1.3 (291) |
| Janssen - 2 vaccines | 0.0 (0) | 0.0 (0) | 0.0 (0) | 0.6 (137) |
| Pfizer - 2 vaccines | 0.0 (1) | 0.2 (15) | 0.1 (16) | 3.0 (670) |

ART: Antiretroviral therapy

By wave (see SDC A), over time, there was an increasingly higher prevalence of diagnosed infections with low recent CD4 cell counts, unsuppressed recent viral load, no previous ART, ongoing tuberculosis, or COPD. In line with the rollout of vaccines, infected persons that had received their primary vaccines increased from 4.1% during wave 3 to 38.9% during wave 4.

### Associations with mortality: 15-39 years old

Estimated from 9 146 first diagnosed infections, the mortality risk was 2.9% (95% CI: 2.6,3.2%). Focusing on person-level characteristics (Figure 2), higher mortality was associated with being older, and with measures indicating worse HIV disease control, i.e., lower recent CD4 (aOR [95% CI]: 3.39 [1.83,6.47] for 0-49 cells/uL and 2.20 [1.23;4.04] for 50-199 cells/uL, each versus ≥500); no evidence of ART collection (2.30 [1.47,3.54]); among those on ART, high or unknown recent VL (1.53 [1.03,2.24] for ≥1000 copies/ml and 1.26 [0.88,1.81] for unknown, versus <1000); and only recent first HIV evidence (1.37 [0.91,2.05] for evidence <1 year ago). For other comorbidities, in order of decreasing magnitudes of associations, those with ongoing tuberculosis, CKD, tuberculosis diagnosed <1 year ago, diabetes, tuberculosis diagnosed ≥1 year ago, or hypertension experienced 2-8 times higher odds of mortality, though there may be an attenuation of the individual condition effects in those with multiple conditions. There were no deaths amongst the 110 infections that occurred in people who had received at least 1 vaccine. Higher admission pressure and later waves were associated with higher mortality (see SDC B for all univariable and multivariable results).

**Figure 2:**
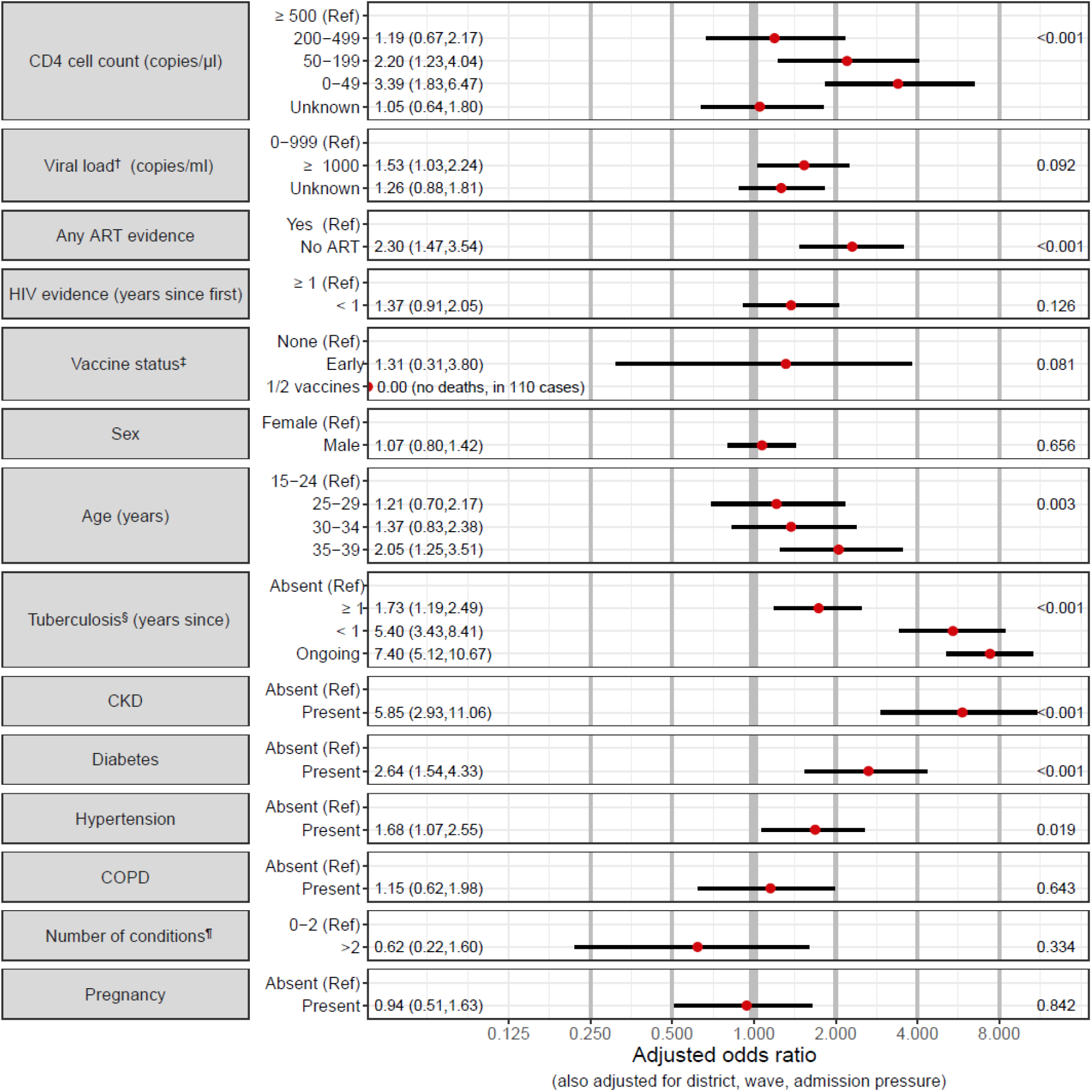
Adjusted odds ratios (and 95% CIs) describing the association of mortality with a person’s HIV characteristics, vaccination, demographic characteristics, and co-existing conditions, for 15–39-year-old adults with HIV and SARS-CoV-2 infections in the Western Cape. P-values are reported on the right. ^†^ Among those with evidence of ART at any time. ^‡^ The highest level of protection from among: No vaccine received (‘None’); <28 days since a first vaccine dose (‘Early’); ≥28 days since (at least) a first dose of a vaccine (1 or 2 Pfizer or Janssen vaccines not distinguished due to small samples). ^§^ Time since the most recent evidence of a start of an episode; with episodes beginning after 2 months before the SARS-CoV-2 diagnosis considered as ongoing. ^¶^ Hypertension, diabetes, chronic obstructive pulmonary disease, chronic kidney disease, tuberculosis (at any time). ART: Antiretroviral therapy. CI: Confidence interval. CKD: Chronic kidney disease. COPD: Chronic obstructive pulmonary disease.

### Associations with mortality: ≥40 years old

Estimated from 8 675 first diagnosed infections, the mortality risk was 8.6% (95% CI: 8.1,9.2%). Older age was most strongly associated with higher mortality with a moderate increase in mortality in men (Figure 3). As in the younger age group, most measures of poor HIV disease control were associated with higher mortality: lower recent CD4 (aOR [95% CI]: 3.30 [2.04,5.31] for 0-49 cells/uL and 1.82 [1.25,2.66] for 50-199, versus ≥500), no ART (1.48 [1.12,1.94]) and having first HIV evidence within the last year (1.48 [1.08,2.01]); though there was no evidence suggesting an association with VL in those on ART. While comorbidities were also strongly associated with mortality, the associations were attenuated compared to those in 15-39-year-olds. Vaccination was strongly protective (0.10 [0.01; 0.47] for the primary series and 0.40 [0.16; 0.89] for 1 dose of the Pfizer vaccine, versus no vaccine).

**Figure 3:**
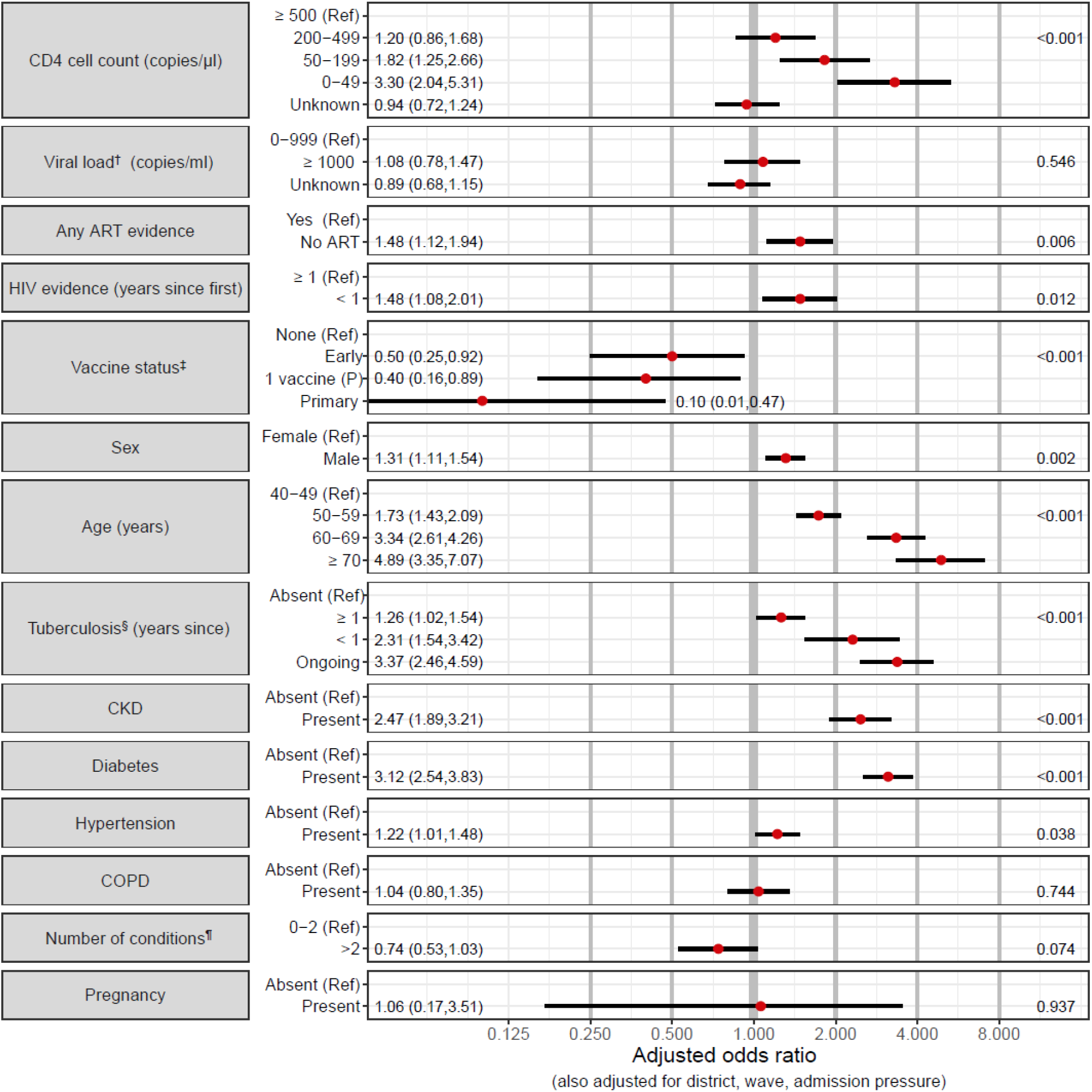
Adjusted odds ratios describing the association of mortality with a person’s HIV characteristics, vaccination, demographic characteristics, and co-existing conditions, for adults with HIV and SARS-CoV-2 infections aged ≥ 40 years in the Western Cape. P-values are reported on the right. ^†^ Among those with evidence of ART at any time. ^‡^ The highest level of protection from among: No vaccine received (‘None’); <28 days since a first vaccine dose (‘Early’); ≥28 days since 1 dose of a Pfizer vaccine (‘1 vaccine (P)’); ≥28 days since 1 Janssen vaccine, possibly with a 2^nd^ vaccine too, or ≥14 days since a 2nd Pfizer vaccine |(combined into the ‘primary’ series due to the small sample). ^§^ Time since the most recent evidence of a start of an episode; with episodes beginning after 2 months before the SARS-CoV-2 diagnosis considered as ongoing. ^¶^ Hypertension, diabetes, chronic obstructive pulmonary disease, chronic kidney disease, tuberculosis (at any time). ART: Antiretroviral therapy. CKD: Chronic kidney disease. COPD: Chronic obstructive pulmonary disease.

### Secondary analyses

The associations of mortality with CD4 were similar (younger age group) or slightly dampened (older age group) when using CD4 nadir instead, while CD4 at ART initiation offered the least predictive power (see SDC C). Overall findings remained similar when including wave 4 infections (see SDC D).

## Discussion

Among more than 17 000 first diagnosed SARS-CoV-2 infections in adult PWH during the ancestral, Beta and Delta variant waves (waves 1 to 3), mortality was strongly associated with suboptimal HIV control indicated by low CD4 cell count, no evidence of starting ART, only recent first HIV evidence, and, in those aged <40 years, VL ≥1000 copies/ml despite ART; and prevalence of these risk factors increased during COVID-19 waves 1 to 4. Mortality was also associated with the well-established risk factors of older age, male sex (in those ≥40 years old), and comorbidities including tuberculosis, which were present in half of those with SARS-CoV-2 infections. Vaccination was strongly protective against death.

To our knowledge, this is the largest study of SARS-CoV-2 infections including non-hospitalized PWH, and one of few studies from sub-Saharan Africa where most PWH live and where a greater proportion of people may be immunosuppressed and/or not on suppressive ART, allowing us to robustly assess their associations with mortality [1, 7, 8]. While our finding that low CD4 cell count is associated with COVID-19 mortality concurs with a large US study [4], we also showed increased mortality if not on ART, or virologically unsuppressed after starting ART for 15-39-year-olds, with stronger associations than those previously shown by the WHO Global Clinical Platform of COVID-19, which was restricted to hospitalized patients [7]. While most people in our cohort were known to be living with HIV for several years, 2% had a first HIV evidence within three weeks of the SARS-CoV-2 diagnosis, suggesting both diagnoses may have occurred in the same health encounter. Importantly, among those aged ≥40 years, mortality was associated with recent first HIV evidence, which, in this age group, could indicate delayed HIV diagnosis or start of ART, and thus suggests delayed HIV care access. The wave pattern observed in our study population (a relatively large first wave) mirrored that in the lower socio-economic status subdistricts in the WC [19, 20], consistent with the burden of HIV in South Africa being disproportionately higher among the poor.

The importance of comorbidities in driving mortality, especially in younger PWH, has been demonstrated in several studies [1, 5-8, 21]. The high prevalence of comorbidities is notable; however, this may be impacted by testing guidelines during COVID-19 waves which restricted testing to older people or those with comorbidities [19]. These testing restrictions may also have biased towards PWH with poorer disease control, since younger virologically suppressed PWH were not eligible for testing during earlier COVID-19 waves. Nonetheless, the high proportion of SARS-CoV-2 infections with previous/ongoing tuberculosis and associated increased risk of mortality confirms our earlier findings and it remains difficult to determine the extent of exacerbation of tuberculosis by SARS-CoV-2 infection and vice versa [3].

Our findings have several important implications for reducing poor SARS-CoV-2 outcomes in PWH. First, it remains a public health priority to ensure all PWH are on suppressive ART, as suboptimal HIV control is a key correlate of SARS-CoV-2 mortality and determinant of prolonged infection with the risk of accumulation of mutations [13, 14]. This is especially important considering the insults to routine health services, including HIV services, during the COVID-19 pandemic [22-24]. Concerningly, we found an increasing and high proportion of patients with CD4 <200 cells/µL and VL ≥1000 copies/ml, possibly reflecting gaps in HIV care earlier in the pandemic. Second, we need to determine and implement the most effective SARS-CoV-2 vaccination strategies for PWH [10]. Third, comorbidity diagnosis and management in PWH should be optimized, with a particular focus on diagnosis and treatment of tuberculosis which has been severely impacted by COVID-19 [25]. Finally, among PWH with COVID-19 diagnoses, those with suboptimal HIV control (preceding low CD4 cell count especially <200 cells/µL, no ART evidence, VL ≥1000 copies/ml) should be considered for anti-SARS-CoV-2 therapy if available, in addition to using the COVID-19 episode as an opportunity to optimize their HIV management.

While strengths of our study include the large size and ability to link to pre-existing HIV care and comorbidity data with ascertainment of ART, VL and CD4 cell count measures that were not reliant on patient recall, it also has several limitations. Routine public sector data may not include ART and other medications dispensed in the private sector and comorbidities are likely under-ascertained due to reliance on algorithmic inference based on public sector laboratory tests, ICD-10 coding and treatments. We were also unable to adjust for comorbidities and behaviours that could not be algorithmically inferred such as obesity, smoking, malnutrition, as well as socio-economic status (though adjusted for district). Because of the substantial under-ascertainment of all SARS-CoV-2 infections, we did not attempt to assess the impact of a prior SARS-CoV-2 infection on a subsequent COVID-19 outcome and restricted our analysis to first diagnosed infections, with a low risk of reinfection during our analysis period which was prior to the emergence of the Omicron variant.

Interpretation of our results is restricted to those with diagnosed SARS-CoV-2 infections, which several seroprevalence studies in the Western Cape and across South Africa have indicated represent only a small fraction of all infections [26]. Compared to the population of all people with diagnosed or undiagnosed SARS-CoV-2 infection, our population will over-represent those with better testing coverage, especially during the wave surges when public sector testing was restricted, which includes older patients, women who are pregnant, and those with more severe HIV, other comorbidities, or severe COVID-19 disease. Increased testing in those with severe COVID-19 disease also likely attenuated the associations of mortality with the indicators of HIV control [27-29]. While we could not distinguish between deaths due to COVID-19 *per se*, and those where the SARS-CoV-2 diagnosis was incidental in a patient with an alternative primary cause of death, to mitigate against this, we restricted our primary analysis to the first three waves and an analysis of deaths in admitted patients showed that the vast majority of patients dying with SARS-COV-2 in this period did have COVID-19 pneumonia [18]. We also showed similar associations of HIV disease severity characteristics with mortality in the analysis including wave 4, and it is reasonable to expect that these characteristics will remain important predictors of mortality due to future variants. Lack of specificity of the outcome of COVID-19 death and other diagnoses such as Pneumocystis jiroveci pneumonia (PJP) in these patients also means that we cannot exclude that some of the association between low CD4 cell count and mortality in patients with COVID-19 may be from pathology due to PJP rather than COVID-19 itself.

## Conclusions

Among PWH and SARS-CoV-2 infections, markers of suboptimal HIV management increased in later waves and are modifiable risk factors for poor COVID-19 outcomes in addition to tuberculosis and other comorbidities, and should be considered when prioritizing patients for anti-SARS-CoV-2 therapy. Maximizing efforts to ensure that PWH are on suppressive ART with optimal comorbidity diagnosis and management as well as SARS-CoV-2 vaccination should be public health priorities for preventing both poor COVID-19 outcomes and poor HIV outcomes.

## Supporting information

Supplemental Digital Content (SDC)

STROBE checklist

## Data Availability

The data are not publicly available due to privacy or ethical restrictions. The data that support the findings of this study can be requested from the Western Cape Provincial Health Data Centre (WCPHDC) [https://www.westerncape.gov.za/general-publication/provincial-health-data-centre]; restrictions apply to the availability of these data.

## Authors’ contributions

MD, AB, NZ, RK and NT conceived the study. NZ prepared the data extracts. RK led the analysis of the data and produced manuscript outputs, with analysis contributions by NZ, ON, ROY, TJ and SB, and critical reviews of the analysis outputs by MD, AB, EM, VT and NT. RK, MD and NZ drafted and finalized the manuscript. All authors reviewed the manuscript, provided inputs and approved the manuscript.

## Acknowledgements

We would like to acknowledge all patients in the Western Cape and to thank the Western Cape Department of Health Provincial Health Data Centre (WCPHDC), the South African National Department of Health and the Electronic Vaccine Data System, the Western Cape Department of Health COVID-19 Outbreak Response Team, the Western Cape Communicable Disease Control sub-directorate and Western Cape health care workers involved in the COVID-19 response for their contributions to this report.

