## Supplemental Digital Content (SDC) for "COVID-19 among adults living with HIV: Correlates of mortality in a general population in a resource-limited setting"

**Additional analysis outputs for:**

| | |
|---|---|
| A: Additional descriptions of SARS-CoV-2 infections | 2 |
| B: Unadjusted and adjusted odds ratios for all model terms | 4 |
| C: Adjusted odds ratios using alternative CD4 cell counts measures | 8 |
| D: Adjusted odds ratios for models including SARS-CoV-2 infections diagnosed during Wave 4 | 12 |

### Supplemental Digital Content A: Additional descriptions of SARS-CoV-2 infections

##### Table A1: Admission pressure, timing and location for SARS-CoV-2 infections among adults with HIV in the Western Cape, stratified by age group (percentages and counts)

| Characteristic | Cases until end of Wave 3 | | | All cases (including Wave 4) |
|---|---|---|---|---|
| | Ages 15-39 years n = 9,321 | Ages ≥40 years n = 8,799 | All ages n = 18,120 | All ages n = 22,057 |
| **Admission pressure[1] (%)** | | | | |
| <25 | 28.7 (2,674) | 23.5 (2,070) | 26.2 (4,744) | 29.2 (6,432) |
| ≥25 and <50 | 34.1 (3,181) | 36.5 (3,208) | 35.3 (6,389) | 37.8 (8,346) |
| ≥50 | 37.2 (3,466) | 40.0 (3,521) | 38.6 (6,987) | 33.0 (7,279) |
| **Wave** | | | | |
| Wave 1 | 32.7 (3,052) | 31.0 (2,731) | 31.9 (5,783) | 26.2 (5,783) |
| Wave 2 | 26.5 (2,469) | 28.8 (2,532) | 27.6 (5,001) | 22.7 (5,001) |
| Wave 3 | 29.3 (2,731) | 31.2 (2,747) | 30.2 (5,478) | 24.8 (5,478) |
| Wave 4 | - | - | - | 15.6 (3,430) |
| Inter-waves | 11.5 (1,069) | 9.0 (789) | 10.3 (1,858) | 10.7 (2,365) |
| **District** | | | | |
| Cape Winelands | 9.8 (915) | 10.8 (950) | 10.3 (1,865) | 10.2 (2,253) |
| Central Karoo | 1.1 (106) | 1.4 (124) | 1.3 (230) | 1.3 (277) |
| City of Cape Town | 61.1 (5,698) | 62.7 (5,518) | 61.9 (11,216) | 62.1 (13,707) |
| Garden Route | 18.0 (1,677) | 16.1 (1,416) | 17.1 (3,093) | 17.2 (3,792) |
| Overberg | 5.2 (489) | 4.3 (379) | 4.8 (868) | 4.6 (1,009) |
| West Coast | 4.7 (436) | 4.7 (412) | 4.7 (848) | 4.6 (1,019) |

[1] The week's total number of COVID-19 admissions relative to the maximum weekly COVID-19 admissions for that district (as a percent).

##### Table A2: Alternative CD4 cell count measures for SARS-CoV-2 infections among adults with HIV in the Western Cape, stratified by age group (percentages and counts)

| CD4 count (cells/µl) measure | Cases until end of Wave 3 | | | All cases (including Wave 4) |
|---|---|---|---|---|
| | Ages 15-39 years n = 9,321 | Ages ≥40 years n = 8,799 | All ages n = 18,120 | All ages n = 22,057 |
| **Recent** | | | | |
| ≥ 500 | 14.0 (1,304) | 11.3 (991) | 12.7 (2,295) | 12.6 (2,775) |
| 200-499 | 15.7 (1,465) | 11.5 (1,014) | 13.7 (2,479) | 13.5 (2,986) |
| 50-199 | 7.0 (648) | 5.8 (511) | 6.4 (1,159) | 7.1 (1,576) |
| 0-49 | 3.6 (333) | 2.2 (191) | 2.9 (524) | 3.5 (773) |
| Unknown | 59.8 (5,571) | 69.2 (6,092) | 64.4 (11,663) | 63.2 (13,947) |
| **Nadir** | | | | |
| ≥ 500 | 15.3 (1,424) | 12.1 (1,067) | 13.7 (2,491) | 13.4 (2,965) |
| 200-499 | 44.1 (4,109) | 39.2 (3,446) | 41.7 (7,555) | 40.7 (8,986) |
| 50-199 | 21.3 (1,987) | 28.7 (2,526) | 24.9 (4,513) | 25.1 (5,535) |
| 0-49 | 8.0 (743) | 10.1 (886) | 9.0 (1,629) | 9.6 (2,112) |
| Unknown | 11.4 (1,058) | 9.9 (874) | 10.7 (1,932) | 11.1 (2,459) |
| **At ART initiation** | | | | |
| ≥ 500 | 11.9 (1,105) | 7.2 (631) | 9.6 (1,736) | 9.6 (2,111) |
| 200-499 | 34.0 (3,166) | 25.5 (2,245) | 29.9 (5,411) | 29.8 (6,572) |
| 50-199 | 14.9 (1,387) | 19.9 (1,752) | 17.3 (3,139) | 17.3 (3,813) |
| 0-49 | 3.8 (358) | 5.4 (477) | 4.6 (835) | 4.7 (1,027) |
| Unknown | 35.5 (3,305) | 42.0 (3,694) | 38.6 (6,999) | 38.7 (8,534) |

**Table A3: Outcomes, demographic characteristics, co-existing conditions, vaccination status and HIV characteristics, for SARS-CoV-2 infections among adults with HIV in the Western Cape, stratified by wave (percentages and counts)**

| Characteristic / Outcome | Wave 1<br>n = 5,783 | Wave 2<br>n = 5,001 | Wave 3<br>n = 5,478 | Wave 4<br>n = 3,430 | Inter-wave<br>n = 2,361 |
|---|---|---|---|---|---|
| **Death** | 5.6 (323) | 5.6 (281) | 5.9 (325) | 4.9 (167) | 5.8 (137) |
| **Sex** | | | | | |
| Female | 75.2 (4,351) | 70.3 (3,515) | 67.2 (3,681) | 68.9 (2,364) | 69.8 (1,650) |
| Male | 24.8 (1,432) | 29.7 (1,486) | 32.8 (1,797) | 31.1 (1,066) | 30.2 (715) |
| **Age (years)** | 0 | 0 | 0 | 0 | 0 |
| 15-24 | 4.8 (279) | 4.1 (204) | 5.1 (277) | 4.7 (161) | 7.8 (185) |
| 25-29 | 11.3 (655) | 10.9 (544) | 10.3 (566) | 12.7 (435) | 13.4 (317) |
| 30-34 | 17.5 (1,012) | 16.6 (830) | 15.9 (872) | 20.3 (695) | 18.3 (432) |
| 35-39 | 19.1 (1,106) | 17.8 (891) | 18.5 (1,016) | 19.5 (669) | 18.9 (448) |
| 40-49 | 28.7 (1,657) | 29.7 (1,486) | 30.2 (1,653) | 26.1 (894) | 26.1 (618) |
| 50-59 | 13.7 (792) | 14.8 (742) | 15.0 (822) | 12.2 (419) | 11.9 (282) |
| 60-69 | 3.9 (225) | 4.7 (233) | 4.0 (217) | 3.7 (127) | 2.6 (62) |
| ≥70 | 1.0 (57) | 1.4 (71) | 1.0 (55) | 0.9 (30) | 0.9 (21) |
| **Hypertension** | 21.2 (1,226) | 21.2 (1,062) | 20.0 (1,096) | 16.3 (559) | 17.6 (417) |
| **Diabetes** | 11.8 (684) | 10.1 (506) | 7.6 (418) | 6.2 (212) | 7.6 (179) |
| **Chronic obstructive pulmonary disease** | 5.9 (343) | 7.7 (384) | 8.1 (442) | 10.6 (365) | 7.6 (180) |
| **Chronic kidney disease** | 4.0 (233) | 3.7 (183) | 3.4 (187) | 2.8 (95) | 3.6 (86) |
| **Tuberculosis[1] (years since)** | | | | | |
| Absent | 72.2 (4,177) | 71.7 (3,586) | 67.5 (3,695) | 60.8 (2,087) | 64.4 (1,523) |
| ≥ 1 | 21.5 (1,242) | 21.3 (1,064) | 23.8 (1,303) | 23.3 (800) | 21.2 (502) |
| < 1 | 3.0 (174) | 2.7 (136) | 3.4 (187) | 5.7 (194) | 4.9 (115) |
| Ongoing | 3.3 (190) | 4.3 (215) | 5.3 (293) | 10.2 (349) | 9.5 (225) |
| **Pregnancy (in women aged 15-49 years)** | 7.1 (263) | 7.5 (216) | 7.0 (215) | 8.1 (165) | 11.7 (169) |
| **Vaccination status[2]** | | | | | |
| None | 100.0 (5,783) | 100.0 (5,001) | 88.8 (4,866) | 54.4 (1,866) | 94.9 (2,245) |
| Early | - | - | 5.5 (298) | 1.7 (57) | 1.0 (24) |
| Janssen - 1 vaccine | - | - | 3.8 (207) | 16.8 (577) | 1.5 (35) |
| Pfizer - 1 vaccine | - | - | 1.7 (91) | 5.1 (174) | 1.1 (26) |
| Janssen - 2 vaccines | - | - | 0.0 (0) | 3.9 (133) | 0.2 (4) |
| Pfizer - 2 vaccines | - | - | 0.3 (16) | 18.2 (623) | 1.3 (31) |
| **Number of previous SARS-CoV-2 diagnoses** | | | | | |
| 0 | 100.0 (5,783) | 98.4 (4,920) | 96.9 (5,308) | 87.8 (3,011) | 97.0 (2,294) |
| 1 | 0.0 (0) | 1.6 (81) | 3.0 (166) | 11.8 (406) | 2.9 (69) |
| 2 | 0.0 (0) | 0.0 (0) | 0.1 (4) | 0.4 (13) | 0.1 (2) |
| **CD4 count (cells/ul)** | | | | | |
| ≥ 500 | 11.6 (670) | 13.7 (684) | 13.3 (729) | 12.7 (437) | 10.8 (255) |
| 200-499 | 13.4 (775) | 12.8 (642) | 14.3 (783) | 12.4 (426) | 15.2 (360) |
| 50-199 | 5.2 (301) | 6.0 (300) | 7.4 (407) | 10.2 (349) | 9.3 (219) |
| 0-49 | 2.4 (140) | 2.6 (132) | 2.8 (152) | 6.1 (210) | 5.9 (139) |
| Unknown | 67.4 (3,897) | 64.8 (3,243) | 62.2 (3,407) | 58.5 (2,008) | 58.9 (1,392) |
| **Viral load[3] (copies/ml)** | | | | | |
| 0-999 | 74.7 (3,970) | 72.7 (3,301) | 69.7 (3,449) | 59.7 (1,833) | 64.4 (1,360) |
| ≥ 1000 | 6.6 (352) | 8.9 (405) | 10.8 (532) | 13.3 (407) | 14.5 (307) |
| Unknown | 18.7 (991) | 18.4 (837) | 19.5 (966) | 27.0 (830) | 21.0 (444) |
| **ART (years since most recent collection)** | | | | | |
| ≥ 2 | 49.6 (2,868) | 41.2 (2,061) | 45.9 (2,517) | 49.9 (1,713) | 41.7 (987) |
| < 2 | 42.3 (2,445) | 49.6 (2,482) | 44.4 (2,430) | 39.6 (1,357) | 47.5 (1,124) |
| No ART | 8.1 (470) | 9.2 (458) | 9.7 (531) | 10.5 (360) | 10.7 (254) |
| **HIV (years since first evidence)** | | | | | |
| <1 | 8.0 (464) | 6.9 (347) | 6.3 (346) | 8.8 (303) | 10.4 (246) |
| 1-5 | 31.7 (1,835) | 32.0 (1,602) | 29.3 (1,607) | 29.3 (1,006) | 32.8 (776) |
| 5-10 | 34.9 (2,016) | 34.8 (1,738) | 37.0 (2,026) | 35.7 (1,224) | 32.9 (777) |
| ≥ 10 | 25.4 (1,468) | 26.3 (1,314) | 27.4 (1,499) | 26.2 (897) | 23.9 (566) |

[1] Time since the most recent evidence of a start of an episode; with episodes beginning after 2 months before the SARS-CoV-2 diagnosis considered as ongoing. [2] The highest level of protection from among six categories: No vaccine received ('None'); <28 days since a first vaccine dose ('Early'); ≥28 days since a first dose of a vaccine, by type ('1 vaccine'); or ≥14 days since a second dose, by type ('2 vaccines'). [3] Among those with evidence of ART at any time.

### Supplemental Digital Content B: Unadjusted and adjusted odds ratios for all model terms

**Table B1: Mortality rates, unadjusted and adjusted odds ratios (and 95% CIs) describing the association of mortality with a person's demographic characteristics, co-existing conditions, vaccination status, HIV characteristics and time/location covariates, for 15–39-year-old adults with HIV and SARS-CoV-2 infections in the Western Cape**

| Variable | Category | n | Mortality risk (95% CI) | Univariable analysis: Unadjusted odds ratio (uOR) | | Multivariable analysis: Adjusted odds ratio (aOR) | |
|---|---|---|---|---|---|---|---|
| | | | | Estimate (95% CI) | p-value | Estimate (95% CI) | p-value |
| **Sex** | **Female (Ref)** | 7114 | 2.52 (2.16,2.91) | - | <0.001 | | 0.656 |
| | **Male** | 2032 | 4.13 (3.31,5.09) | 1.67 (1.28,2.17) | | 1.07 (0.80,1.42) | |
| **Age group (years)** | **15-24 (Ref)** | 884 | 2.26 (1.39,3.47) | - | 0.001 | | 0.003 |
| | **25-29** | 1973 | 2.13 (1.54,2.87) | 0.94 (0.56,1.64) | | 1.21 (0.70,2.17) | |
| | **30-34** | 3008 | 2.56 (2.03,3.19) | 1.13 (0.70,1.92) | | 1.37 (0.83,2.38) | |
| | **35-39** | 3281 | 3.78 (3.15,4.49) | 1.70 (1.08,2.82) | | 2.05 (1.25,3.51) | |
| **Hypertension** | **Absent (Ref)** | 8360 | 2.75 (2.41,3.12) | - | 0.029 | | 0.019 |
| | **Present** | 786 | 4.20 (2.91,5.85) | 1.55 (1.05,2.22) | | 1.68 (1.07,2.55) | |
| **Diabetes** | **Absent (Ref)** | 8789 | 2.74 (2.41,3.11) | - | 0.001 | | <0.001 |
| | **Present** | 357 | 6.16 (3.90,9.18) | 2.33 (1.45,3.57) | | 2.64 (1.54,4.33) | |
| **Chronic obstructive pulmonary disease** | **Absent (Ref)** | 8776 | 2.83 (2.49,3.19) | - | 0.19 | | 0.643 |
| | **Present** | 370 | 4.05 (2.29,6.60) | 1.45 (0.82,2.39) | | 1.15 (0.62,1.98) | |
| **Chronic kidney disease** | **Absent (Ref)** | 9062 | 2.74 (2.41,3.09) | - | <0.001 | | <0.001 |
| | **Present** | 84 | 17.86 (10.35,27.74) | 7.73 (4.20,13.32) | | 5.85 (2.93,11.06) | |
| **Tuberculosis[1] (years since)** | **Absent (Ref)** | 6708 | 1.46 (1.19,1.78) | - | <0.001 | | <0.001 |
| | **≥ 1** | 1644 | 2.98 (2.21,3.92) | 2.07 (1.45,2.92) | | 1.73 (1.19,2.49) | |
| | **< 1** | 319 | 13.17 (9.66,17.38) | 10.23 (6.93,14.87) | | 5.40 (3.43,8.41) | |
| | **Recent/current** | 475 | 15.58 (12.44,19.16) | 12.45 (9.03,17.09) | | 7.40 (5.12,10.67) | |
| **Number of conditions[2]** | **0-2 (Ref)** | 9076 | 2.81 (2.48,3.17) | - | 0.001 | | 0.334 |
| | **>2** | 70 | 11.43 (5.07,21.28) | 4.46 (1.96,8.88) | | 0.62 (0.22,1.60) | |
| **Pregnancy** | **Absent (Ref)** | 8381 | 2.97 (2.62,3.36) | - | 0.054 | | 0.842 |
| | **Present** | 765 | 1.83 (1.00,3.05) | 0.61 (0.34,1.01) | | 0.94 (0.51,1.63) | |

Continued on next page

**Table B1 continued**

| Variable | Category | n | Mortality risk (95% CI) | Univariable analysis: Unadjusted odds ratio (uOR) Estimate (95% CI) | p-value | Multivariable analysis: Adjusted odds ratio (aOR) Estimate (95% CI) | p-value |
|---|---|---|---|---|---|---|---|
| **Vaccination status**[3] | **None (Ref)** | 8967 | 2.90 (2.56,3.27) | | 0.032 | | 0.081 |
| | **Early** | 69 | 4.35 (0.91,12.18) | 1.52 (0.37,4.13) | | 1.31 (0.31,3.80) | |
| | **1/2 vaccines** | 110 | 0.00 (0.00,3.30) | 0.00 (no deaths) | | 0.00 (0.00,0.29) | |
| **CD4 count (cells/µl)** | **≥ 500 (Ref)** | 1280 | 1.48 (0.90,2.31) | - | <0.001 | | <0.001 |
| | **200-499** | 1441 | 2.50 (1.76,3.44) | 1.70 (0.98,3.04) | | 1.19 (0.67,2.17) | |
| | **50-199** | 631 | 7.77 (5.80,10.14) | 5.59 (3.32,9.80) | | 2.20 (1.23,4.04) | |
| | **0-49** | 318 | 14.78 (11.07,19.17) | 11.51 (6.76,20.38) | | 3.39 (1.83,6.47) | |
| | **Unknown** | 5476 | 2.05 (1.69,2.46) | 1.39 (0.87,2.33) | | 1.05 (0.64,1.80) | |
| **Viral load**[4] **(copies/ml)** | **0-999 (Ref)** | 5546 | 1.77 (1.44,2.15) | - | <0.001 | | 0.092 |
| | **≥ 1000** | 892 | 7.06 (5.47,8.95) | 4.22 (3.04,5.83) | | 1.53 (1.03,2.24) | |
| | **Unknown** | 1896 | 3.01 (2.28,3.88) | 1.72 (1.23,2.39) | | 1.26 (0.88,1.81) | |
| **ART collection** | **Yes** | 8334 | 2.62 (2.28,2.98) | - | <0.001 | | <0.001 |
| | **No ART** | 812 | 5.54 (4.07,7.35) | 2.18 (1.55,3.01) | | 2.30 (1.47,3.54) | |
| **HIV (years since first evidence)** | **≥ 1 (Ref)** | 8322 | 2.66 (2.32,3.02) | - | <0.001 | | 0.126 |
| | **< 1** | 824 | 5.10 (3.70,6.83) | 1.97 (1.39,2.73) | | 1.37 (0.91,2.05) | |
| **Admission pressure**[5] **(%)** | **<25 (Ref)** | 2630 | 2.78 (2.18,3.48) | - | 0.3 | | 0.036 |
| | **≥25 and <50** | 3147 | 3.24 (2.65,3.92) | 1.17 (0.87,1.60) | | 1.65 (1.11,2.49) | |
| | **≥50** | 3369 | 2.61 (2.10,3.21) | 0.94 (0.69,1.29) | | 1.24 (0.79,2.00) | |
| **Wave** | **Wave 1 (Ref)** | 3052 | 2.23 (1.73,2.82) | - | 0.008 | | 0.022 |
| | **Wave 2** | 2415 | 2.61 (2.01,3.33) | 1.18 (0.83,1.66) | | 1.29 (0.82,2.01) | |
| | **Wave 3** | 2636 | 3.57 (2.89,4.35) | 1.62 (1.18,2.23) | | 1.68 (1.13,2.50) | |
| | **Inter-waves** | 1043 | 3.64 (2.59,4.97) | 1.66 (1.10,2.47) | | 1.83 (1.08,3.11) | |
| **District** | **City of Cape Town (Ref)** | 5599 | 2.82 (2.40,3.29) | - | 0.209 | | 0.136 |
| | **Cape Winelands** | 907 | 4.08 (2.89,5.58) | 1.46 (1.00,2.09) | | 1.70 (1.12,2.52) | |
| | **Central Karoo** | 104 | 1.92 (0.23,6.77) | 0.68 (0.11,2.16) | | 0.50 (0.08,1.75) | |
| | **Garden Route** | 1623 | 2.40 (1.71,3.27) | 0.85 (0.59,1.20) | | 0.95 (0.64,1.37) | |
| | **Overberg** | 483 | 3.52 (2.06,5.58) | 1.26 (0.73,2.03) | | 1.37 (0.76,2.32) | |
| | **West Coast** | 430 | 2.33 (1.12,4.24) | 0.82 (0.40,1.49) | | 0.94 (0.45,1.75) | |

[1] Time since the most recent evidence of a start of an episode; with episodes beginning after 2 months before the SARS-CoV-2 diagnosis considered as ongoing. [2] Hypertension, diabetes, chronic obstructive pulmonary disease, chronic kidney disease, tuberculosis (at any time). [3] The highest level of protection from among: No vaccine received ('None'); <28 days since a first vaccine dose ('Early'); ≥28 days since (at least) a first dose of a vaccine (1 or 2 Pfizer or Janssen vaccines not distinguished due to small samples). [4] Among those with evidence of ART at any time. [5] The week's total number of COVID-19 admissions relative to the maximum weekly COVID-19 admissions for that district (as a percent).
ART: Antiretroviral therapy. CI: Confidence interval.

**Table B2: Mortality rates, unadjusted and adjusted odds ratios (and 95% CIs) describing the association of mortality with a person's demographic characteristics, co-existing conditions, vaccination status, HIV characteristics and time/location covariates, for adults with HIV and SARS-CoV-2 infections ≥ 40 years old in the Western Cape**

| Variable | Category | n | Mortality risk (95% CI) | Univariable analysis: Unadjusted odds ratio (uOR) | | Multivariable analysis: Adjusted odds ratio (aOR) | |
|---|---|---|---|---|---|---|---|
| | | | | Estimate (95% CI) | p-value | Estimate (95% CI) | p-value |
| **Sex** | **Female (Ref)** | 5529 | 7.23 (6.57,7.95) | - | <0.001 | | 0.002 |
| | **Male** | 3146 | 11.09 (10.02,12.24) | 1.60 (1.38,1.86) | | 1.31 (1.11,1.54) | |
| **Age group (years)** | **40-49 (Ref)** | 5207 | 5.36 (4.76,6.00) | - | <0.001 | | <0.001 |
| | **50-59** | 2560 | 10.35 (9.20,11.60) | 2.04 (1.71,2.43) | | 1.73 (1.43,2.09) | |
| | **60-69** | 711 | 20.68 (17.75,23.84) | 4.60 (3.70,5.72) | | 3.34 (2.61,4.26) | |
| | **≥ 70** | 197 | 29.44 (23.18,36.34) | 7.37 (5.27,10.20) | | 4.89 (3.35,7.07) | |
| **Hypertension** | **Absent (Ref)** | 5796 | 6.82 (6.18,7.49) | - | <0.001 | | 0.039 |
| | **Present** | 2879 | 12.30 (11.12,13.55) | 1.92 (1.65,2.23) | | 1.22 (1.01,1.48) | |
| **Diabetes** | **Absent (Ref)** | 7298 | 6.48 (5.93,7.07) | - | <0.001 | | <0.001 |
| | **Present** | 1377 | 20.04 (17.96,22.26) | 3.62 (3.08,4.25) | | 3.12 (2.54,3.83) | |
| **Chronic obstructive pulmonary disease** | **Absent (Ref)** | 7782 | 8.30 (7.70,8.94) | - | 0.002 | | 0.744 |
| | **Present** | 893 | 11.53 (9.51,13.81) | 1.44 (1.15,1.79) | | 1.04 (0.80,1.35) | |
| **Chronic kidney disease** | **Absent (Ref)** | 8098 | 7.42 (6.86,8.01) | - | <0.001 | | <0.001 |
| | **Present** | 577 | 25.65 (22.13,29.42) | 4.30 (3.50,5.27) | | 2.47 (1.89,3.21) | |
| **Tuberculosis[1] (years since)** | **Absent (Ref)** | 5799 | 7.23 (6.57,7.92) | - | <0.001 | | <0.001 |
| | **≥ 1** | 2284 | 8.89 (7.75,10.13) | 1.25 (1.05,1.49) | | 1.26 (1.02,1.54) | |
| | **< 1** | 228 | 18.86 (14.00,24.55) | 2.98 (2.09,4.18) | | 2.31 (1.54,3.42) | |
| | **Recent/current** | 364 | 23.08 (18.85,27.75) | 3.85 (2.95,4.99) | | 3.37 (2.46,4.59) | |
| **Number of conditions[2]** | **0-2 (Ref)** | 7994 | 7.36 (6.79,7.95) | - | <0.001 | | 0.074 |
| | **>2** | 681 | 23.64 (20.50,27.02) | 3.90 (3.20,4.73) | | 0.74 (0.53,1.03) | |
| **Pregnancy** | **Absent (Ref)** | 8625 | 8.66 (8.08,9.27) | - | 0.195 | | 0.937 |
| | **Present** | 50 | 4.00 (0.49,13.71) | 0.44 (0.07,1.42) | | 1.06 (0.17,3.51) | |

Continued on next page

**Table B2 continued**

| Variable | Category | n | Mortality risk (95% CI) | Univariable analysis: Unadjusted odds ratio (uOR) Estimate (95% CI) | p-value | Multivariable analysis: Adjusted odds ratio (aOR) Estimate (95% CI) | p-value |
|---|---|---|---|---|---|---|---|
| **Vaccination status[3]** | **None (Ref)** | 8275 | 8.82 (8.22,9.45) | - | 0.001 | | <0.001 |
| | **Early** | 217 | 5.07 (2.56,8.89) | 0.55 (0.28,0.97) | | 0.50 (0.25,0.92) | |
| | **Pfizer - 1 vaccine** | 79 | 8.86 (3.64,17.41) | 1.00 (0.42,2.04) | | 0.40 (0.16,0.89) | |
| | **Primary** | 104 | 0.96 (0.02,5.24) | 0.10 (0.01,0.45) | | 0.10 (0.01,0.47) | |
| **CD4 count (cells/µl)** | **≥ 500 (Ref)** | 974 | 7.80 (6.20,9.67) | - | <0.001 | | <0.001 |
| | **200-499** | 998 | 10.12 (8.32,12.16) | 1.33 (0.98,1.82) | | 1.20 (0.86,1.68) | |
| | **50-199** | 502 | 14.74 (11.76,18.15) | 2.04 (1.45,2.87) | | 1.82 (1.25,2.66) | |
| | **0-49** | 184 | 24.46 (18.43,31.32) | 3.83 (2.53,5.74) | | 3.30 (2.04,5.31) | |
| | **Unknown** | 6017 | 7.53 (6.87,8.22) | 0.96 (0.75,1.25) | | 0.94 (0.72,1.24) | |
| **Viral load[4] (copies/ml)** | **0-999 (Ref)** | 6112 | 7.66 (7.00,8.35) | - | <0.001 | | 0.546 |
| | **≥ 1000** | 570 | 12.63 (10.02,15.64) | 1.74 (1.33,2.26) | | 1.08 (0.78,1.47) | |
| | **Unknown** | 1183 | 7.27 (5.86,8.90) | 0.95 (0.74,1.19) | | 0.89 (0.68,1.15) | |
| **ART collection** | **Yes** | 7865 | 7.96 (7.37,8.58) | - | <0.001 | | 0.006 |
| | **No ART** | 810 | 15.19 (12.78,17.84) | 2.07 (1.67,2.54) | | 1.48 (1.12,1.94) | |
| **HIV (years since first evidence)** | **≥ 1 (Ref)** | 8164 | 8.06 (7.48,8.67) | - | <0.001 | | 0.012 |
| | **< 1** | 511 | 17.81 (14.59,21.41) | 2.47 (1.93,3.13) | | 1.48 (1.08,2.01) | |
| **Admission pressure[5] (%)** | **<25 (Ref)** | 2036 | 8.25 (7.09,9.53) | - | 0.137 | | 0.932 |
| | **≥25 and <50** | 3183 | 9.43 (8.43,10.49) | 1.16 (0.95,1.41) | | 0.99 (0.78,1.27) | |
| | **≥50** | 3456 | 8.13 (7.24,9.09) | 0.98 (0.81,1.20) | | 0.95 (0.72,1.26) | |
| **Wave** | **Wave 1 (Ref)** | 2731 | 9.34 (8.27,10.49) | - | 0.286 | | 0.104 |
| | **Wave 2** | 2505 | 8.58 (7.51,9.75) | 0.91 (0.75,1.10) | | 0.96 (0.74,1.25) | |
| | **Wave 3** | 2672 | 8.35 (7.32,9.46) | 0.88 (0.73,1.07) | | 1.14 (0.89,1.46) | |
| | **Inter-waves** | 767 | 7.30 (5.56,9.38) | 0.76 (0.56,1.03) | | 0.74 (0.51,1.06) | |
| **District** | **City of Cape Town (Ref)** | 5447 | 9.42 (8.66,10.22) | - | 0.002 | | 0.231 |
| | **Cape Winelands** | 941 | 9.03 (7.28,11.05) | 0.96 (0.75,1.21) | | 0.97 (0.74,1.26) | |
| | **Central Karoo** | 121 | 8.26 (4.03,14.67) | 0.87 (0.42,1.58) | | 0.75 (0.35,1.47) | |
| | **Garden Route** | 1388 | 6.56 (5.31,7.99) | 0.67 (0.53,0.85) | | 0.82 (0.64,1.05) | |
| | **Overberg** | 368 | 5.71 (3.57,8.59) | 0.58 (0.36,0.89) | | 0.65 (0.39,1.03) | |
| | **West Coast** | 410 | 7.07 (4.79,10.00) | 0.73 (0.49,1.06) | | 0.77 (0.50,1.14) | |

[1] Time since the most recent evidence of a start of an episode; with episodes beginning after 2 months before the SARS-CoV-2 diagnosis considered as ongoing. [2] Hypertension, diabetes, chronic obstructive pulmonary disease, chronic kidney disease, tuberculosis (at any time). [3] The highest level of protection from among: No vaccine received ('None'); <28 days since a first vaccine dose ('Early'); ≥28 days since 1 dose of a Pfizer vaccine ('1 vaccine (P)'); ≥28 days since 1 Janssen vaccine, possibly with a 2nd vaccine too, or ≥14 days since a 2nd Pfizer vaccine |(combined into the 'primary' series due to the small sample). [4] Among those with evidence of ART at any time. [5] The week's total number of COVID-19 admissions relative to the maximum weekly COVID-19 admissions for that district (as a percent).
ART: Antiretroviral therapy. CI: Confidence interval.

### Supplemental Digital Content C: Adjusted odds ratios using alternative CD4 cell counts measures

**Table C1: Adjusted odds ratios (and 95% CIs) describing the association of mortality with a person's demographic characteristics, co-existing conditions, vaccination status, HIV characteristics and time/location covariates, using alternative CD4 cell count measures for 15–39-year-old adults with HIV and SARS-CoV-2 infections in the Western Cape**

| Variable | Category | Adjusted odds ratio (aOR) | | |
|---|---|---|---|---|
| | | Model 1: Recent CD4 (AIC[6]: 2051) | Model 2: CD4 nadir (AIC: 2043) | Model 3: CD4 at ART start (AIC: 2062) |
| Sex | Female (Ref) | | | |
| | Male | 1.07 (0.80,1.42) | 1.04 (0.78,1.39) | 1.09 (0.81,1.45) |
| Age group (years) | 15-24 (Ref) | | | |
| | 25-29 | 1.21 (0.70,2.17) | 1.15 (0.66,2.06) | 1.14 (0.66,2.03) |
| | 30-34 | 1.37 (0.83,2.38) | 1.30 (0.78,2.24) | 1.28 (0.78,2.22) |
| | 35-39 | 2.05 (1.25,3.51) | 1.88 (1.15,3.21) | 1.91 (1.17,3.27) |
| Hypertension | Absent (Ref) | | | |
| | Present | 1.68 (1.07,2.55) | 1.70 (1.08,2.59) | 1.67 (1.06,2.53) |
| Diabetes | Absent (Ref) | | | |
| | Present | 2.64 (1.54,4.33) | 2.56 (1.49,4.20) | 2.46 (1.44,4.01) |
| Chronic obstructive pulmonary disease | Absent (Ref) | | | |
| | Present | 1.15 (0.62,1.98) | 1.14 (0.62,1.97) | 1.18 (0.64,2.03) |
| Chronic kidney disease | Absent (Ref) | | | |
| | Present | 5.85 (2.93,11.06) | 5.99 (3.01,11.31) | 5.79 (2.90,10.95) |
| Tuberculosis[1] (years since) | Absent (Ref) | | | |
| | ≥ 1 | 1.73 (1.19,2.49) | 1.50 (1.01,2.20) | 1.67 (1.14,2.42) |
| | < 1 | 5.40 (3.43,8.41) | 5.81 (3.71,8.99) | 7.48 (4.88,11.30) |
| | Ongoing | 7.40 (5.12,10.67) | 7.58 (5.24,10.96) | 8.66 (6.06,12.36) |
| Number of conditions[2] | 0-2 (Ref) | | | |
| | >2 | 0.62 (0.22,1.60) | 0.61 (0.22,1.58) | 0.64 (0.23,1.65) |
| Pregnancy | Absent (Ref) | | | |
| | Present | 0.94 (0.51,1.63) | 0.95 (0.51,1.64) | 0.98 (0.53,1.68) |
| Vaccination status[3] | None (Ref) | | | |
| | Early | 1.31 (0.31,3.80) | 1.29 (0.30,3.71) | 1.18 (0.28,3.46) |
| | 1/2 vaccines | 0.00 (0.00,0.29) | 0.00 (0.00,0.00) | 0.00 (0.00,0.00) |
| CD4 count (cells/µl) | ≥ 500 (Ref) | | | |
| | 200-499 | 1.19 (0.67,2.17) | 1.08 (0.67,1.84) | 0.90 (0.55,1.52) |
| | 50-199 | 2.20 (1.23,4.04) | 1.29 (0.76,2.25) | 1.41 (0.84,2.43) |
| | 0-49 | 3.39 (1.83,6.47) | 3.36 (1.95,5.98) | 1.60 (0.84,3.05) |
| | Unknown | 1.05 (0.64,1.80) | 1.16 (0.63,2.17) | 0.63 (0.37,1.11) |
| Viral load[4] (copies/ml) | 0-999 (Ref) | | | |
| | ≥ 1000 | 1.53 (1.03,2.24) | 1.68 (1.15,2.43) | 2.11 (1.46,3.02) |
| | Unknown | 1.26 (0.88,1.81) | 1.35 (0.94,1.93) | 1.43 (1.00,2.04) |
| ART collection | Yes | | | |
| | No ART | 2.30 (1.47,3.54) | 2.33 (1.45,3.66) | 3.32 (1.97,5.62) |
| HIV (years since first evidence) | ≥ 1 (Ref) | | | |
| | < 1 | 1.37 (0.91,2.05) | 1.48 (0.96,2.24) | 1.33 (0.88,1.99) |

Continued on next page

**Table C1 continued**

| Variable | Category | Adjusted odds ratio (aOR) | | |
|---|---|---|---|---|
| | | Model 1: Recent CD4 (AIC[6]: 2051) | Model 2: CD4 nadir (AIC: 2043) | Model 3: CD4 at ART start (AIC: 2062) |
| Admission pressure[5] (%) | <25 (Ref) | | | |
| | ≥25 and <50 | 1.65 (1.11,2.49) | 1.68 (1.13,2.53) | 1.66 (1.12,2.51) |
| | ≥50 | 1.24 (0.79,2.00) | 1.30 (0.82,2.09) | 1.25 (0.80,2.01) |
| Wave | Wave 1 (Ref) | | | |
| | Wave 2 | 1.29 (0.82,2.01) | 1.27 (0.81,1.99) | 1.31 (0.83,2.04) |
| | Wave 3 | 1.68 (1.13,2.50) | 1.71 (1.15,2.54) | 1.66 (1.11,2.47) |
| | Inter-waves | 1.83 (1.08,3.11) | 1.81 (1.07,3.07) | 1.88 (1.11,3.19) |
| District | City of Cape Town (Ref) | | | |
| | Cape Winelands | 1.70 (1.12,2.52) | 1.70 (1.12,2.54) | 1.71 (1.12,2.54) |
| | Central Karoo | 0.50 (0.08,1.75) | 0.55 (0.09,1.88) | 0.51 (0.08,1.77) |
| | Garden Route | 0.95 (0.64,1.37) | 0.94 (0.64,1.37) | 0.96 (0.65,1.39) |
| | Overberg | 1.37 (0.76,2.32) | 1.38 (0.77,2.34) | 1.31 (0.73,2.21) |
| | West Coast | 0.94 (0.45,1.75) | 0.93 (0.45,1.73) | 0.96 (0.46,1.80) |

[1] Time since the most recent evidence of a start of an episode; with episodes beginning after 2 months before the SARS-CoV-2 diagnosis considered as ongoing. [2] Hypertension, diabetes, chronic obstructive pulmonary disease, chronic kidney disease, tuberculosis (at any time). [3] The highest level of protection from among: No vaccine received ('None'); <28 days since a first vaccine dose ('Early'); ≥28 days since (at least) a first dose of a vaccine (1 or 2 Pfizer or Janssen vaccines not distinguished due to small samples). [4] Among those with evidence of ART at any time. [5] The week's total number of COVID-19 admissions relative to the maximum weekly COVID-19 admissions for that district (as a percent). [6] Akaike Information Criterion (AIC) provides relative measure of models' predictive powers; lower AIC values are more favourable.
ART: Antiretroviral therapy. CI: Confidence interval.

**Table C2: Adjusted odds ratios (and 95% CIs) describing the association of mortality with a person's demographic characteristics, co-existing conditions, vaccination status, HIV characteristics and time/location covariates, using alternative CD4 cell count measures, for adults with HIV and SARS-CoV-2 infections ≥ 40 years old in the Western Cape**

| Variable | Category | Adjusted odds ratio (aOR) | | |
|---|---|---|---|---|
| | | Model 1: Recent CD4 (AIC[6]: 4465) | Model 2: CD4 nadir (AIC: 4470) | Model 3: CD4 at ART start (AIC: 4489) |
| Sex | Female (Ref) | | | |
| | Male | 1.31 (1.11,1.54) | 1.30 (1.10,1.53) | 1.33 (1.13,1.57) |
| Age group (years) | 40-49 (Ref) | | | |
| | 50-59 | 1.73 (1.43,2.09) | 1.71 (1.42,2.06) | 1.67 (1.39,2.02) |
| | 60-69 | 3.34 (2.61,4.26) | 3.44 (2.68,4.39) | 3.27 (2.56,4.17) |
| | 70+ | 4.89 (3.35,7.07) | 4.99 (3.41,7.24) | 4.62 (3.16,6.68) |
| Hypertension | Absent (Ref) | | | |
| | Present | 1.22 (1.01,1.48) | 1.23 (1.02,1.49) | 1.22 (1.01,1.48) |
| Diabetes | Absent (Ref) | | | |
| | Present | 3.12 (2.54,3.83) | 3.14 (2.56,3.86) | 3.04 (2.48,3.73) |
| Chronic obstructive pulmonary disease | Absent (Ref) | | | |
| | Present | 1.04 (0.80,1.35) | 1.09 (0.84,1.41) | 1.09 (0.84,1.41) |
| Chronic kidney disease | Absent (Ref) | | | |
| | Present | 2.47 (1.89,3.21) | 2.49 (1.91,3.23) | 2.52 (1.93,3.27) |
| Tuberculosis[1] (years since) | Absent (Ref) | | | |
| | 12m+ | 1.26 (1.02,1.54) | 1.18 (0.96,1.45) | 1.24 (1.01,1.52) |
| | <12m | 2.31 (1.54,3.42) | 2.68 (1.80,3.93) | 2.93 (1.96,4.28) |
| | Recent/current | 3.37 (2.46,4.59) | 3.66 (2.69,4.96) | 4.07 (2.99,5.49) |
| Number of conditions[2] | 0-2 (Ref) | | | |
| | >2 | 0.74 (0.53,1.03) | 0.75 (0.54,1.04) | 0.75 (0.54,1.04) |
| Pregnancy | Absent (Ref) | | | |
| | Present | 1.06 (0.17,3.51) | 1.14 (0.18,3.74) | 1.11 (0.18,3.65) |
| Vaccination status[3] | None (Ref) | | | |
| | Early | 0.50 (0.25,0.92) | 0.49 (0.24,0.90) | 0.49 (0.24,0.90) |
| | Pfizer - 1 vaccine | 0.40 (0.16,0.89) | 0.41 (0.16,0.89) | 0.41 (0.16,0.90) |
| | Primary | 0.10 (0.01,0.47) | 0.10 (0.01,0.48) | 0.09 (0.01,0.44) |
| CD4 count (cells/µl) | ≥ 500 (Ref) | | | |
| | 200-499 | 1.20 (0.86,1.68) | 1.37 (1.02,1.86) | 1.07 (0.74,1.57) |
| | 50-199 | 1.82 (1.25,2.66) | 1.50 (1.11,2.07) | 1.37 (0.95,2.01) |
| | 0-49 | 3.30 (2.04,5.31) | 2.36 (1.65,3.38) | 2.11 (1.36,3.32) |
| | Unknown | 0.94 (0.72,1.24) | 0.79 (0.53,1.19) | 1.21 (0.85,1.76) |
| Viral load[4] (copies/ml) | 0-999 (Ref) | | | |
| | ≥ 1000 | 1.08 (0.78,1.47) | 1.34 (0.99,1.79) | 1.45 (1.08,1.93) |
| | Unknown | 0.89 (0.68,1.15) | 1.03 (0.79,1.34) | 0.94 (0.73,1.21) |
| ART collection | Yes | | | |
| | No ART | 1.48 (1.12,1.94) | 1.80 (1.32,2.43) | 1.40 (1.03,1.88) |
| HIV (years since first evidence) | ≥ 1 (Ref) | | | |
| | < 1 | 1.48 (1.08,2.01) | 1.87 (1.35,2.57) | 1.57 (1.15,2.14) |

Continued on next page

**Table C2 continued**

| Variable | Category | Adjusted odds ratio (aOR) | | |
|---|---|---|---|---|
| | | Model 1: Recent CD4 (AIC[6]: 4465) | Model 2: CD4 nadir (AIC: 4470) | Model 3: CD4 at ART start (AIC: 4489) |
| **Admission pressure[5] (%)** | **<25 (Ref)** | | | |
| | **≥25 and <50** | 0.99 (0.78,1.27) | 0.98 (0.77,1.26) | 1.00 (0.78,1.28) |
| | **≥50** | 0.95 (0.72,1.26) | 0.95 (0.72,1.26) | 0.95 (0.72,1.26) |
| **Wave** | **Wave 1 (Ref)** | | | |
| | **Wave 2** | 0.96 (0.74,1.25) | 0.97 (0.74,1.26) | 0.98 (0.75,1.27) |
| | **Wave 3** | 1.14 (0.89,1.46) | 1.15 (0.90,1.48) | 1.18 (0.91,1.51) |
| | **Inter-waves** | 0.74 (0.51,1.06) | 0.72 (0.50,1.04) | 0.75 (0.52,1.09) |
| **District** | **City of Cape Town (Ref)** | | | |
| | **Cape Winelands** | 0.97 (0.74,1.26) | 1.01 (0.77,1.31) | 1.00 (0.76,1.30) |
| | **Central Karoo** | 0.75 (0.35,1.47) | 0.81 (0.37,1.57) | 0.79 (0.36,1.54) |
| | **Garden Route** | 0.82 (0.64,1.05) | 0.84 (0.65,1.07) | 0.82 (0.64,1.05) |
| | **Overberg** | 0.65 (0.39,1.03) | 0.66 (0.40,1.05) | 0.67 (0.40,1.06) |
| | **West Coast** | 0.77 (0.50,1.14) | 0.79 (0.52,1.18) | 0.80 (0.52,1.18) |

[1] Time since the most recent evidence of a start of an episode; with episodes beginning after 2 months before the SARS-CoV-2 diagnosis considered as ongoing. [2] Hypertension, diabetes, chronic obstructive pulmonary disease, chronic kidney disease, tuberculosis (at any time). [3] The highest level of protection from among: No vaccine received ('None'); <28 days since a first vaccine dose ('Early'); ≥28 days since 1 dose of a Pfizer vaccine ('1 vaccine (P)'); ≥28 days since 1 Janssen vaccine, possibly with a 2nd vaccine too, or ≥14 days since a 2nd Pfizer vaccine |(combined into the 'primary' series due to the small sample). [4] Among those with evidence of ART at any time. [5] The week's total number of COVID-19 admissions relative to the maximum weekly COVID-19 admissions for that district (as a percent). [6] Akaike Information Criterion (AIC) provides relative measure of models' predictive powers; lower AIC values are more favourable.
ART: Antiretroviral therapy. CI: Confidence interval.

### Supplemental Digital Content D: Adjusted odds ratios for models including SARS-CoV-2 infections diagnosed during Wave 4

**Figure D1: Adjusted odds ratios (and 95% CIs) describing the association of mortality with a person's HIV characteristics, vaccination, demographic characteristics, and co-existing conditions, for 15–39-year-old adults with HIV and SARS-CoV-2 infections in the Western Cape using all infections until 10 March 2022.** P-values are reported on the right.

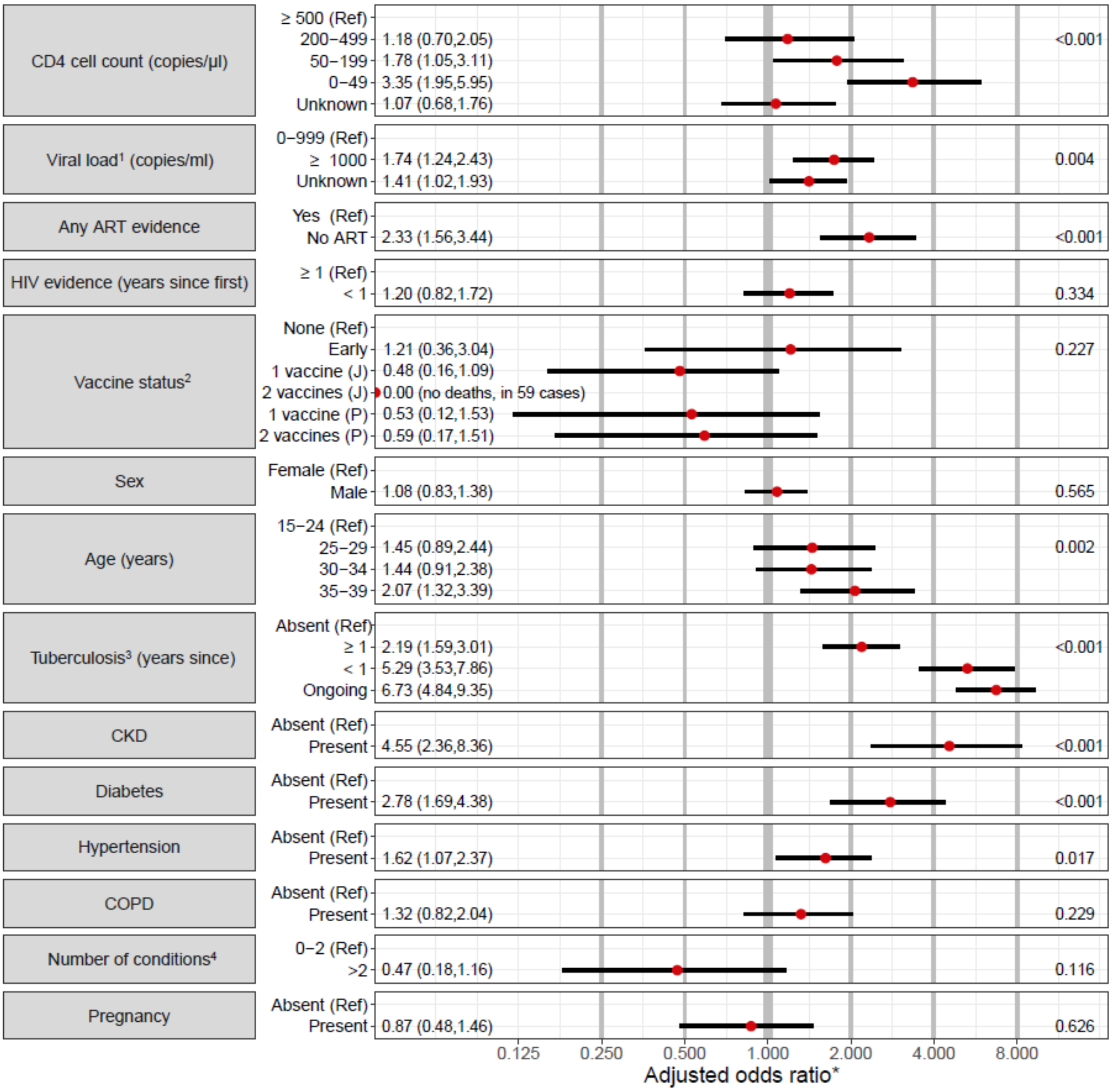


* Also adjusted for district, wave, admission pressure

[1] Among those with evidence of ART at any time. [2] The highest level of protection from among six categories: No vaccine received ('None'); <28 days since a first vaccine dose ('Early'); ≥28 days since a first dose of a vaccine, by type ('1 vaccine'); or ≥14 days since a second dose, by type ('2 vaccines'). J: Janssen, P: Pfizer. [3] Time since the most recent evidence of a start of an episode; with episodes beginning after 2 months before the SARS-CoV-2 diagnosis considered as ongoing. [4] Hypertension, diabetes, chronic obstructive pulmonary disease, chronic kidney disease, tuberculosis (at any time).
ART: Antiretroviral therapy. CI: Confidence interval. CKD: Chronic kidney disease. COPD: Chronic obstructive pulmonary disease.

**Figure D2: Adjusted odds ratios (and 95% CIs) describing the association of mortality with a person's HIV characteristics, vaccination, demographic characteristics, and co-existing conditions, for adults with HIV and SARS-CoV-2 infections aged ≥ 40 years in the Western Cape using all infections until 10 March 2022.** P-values are reported on the right.

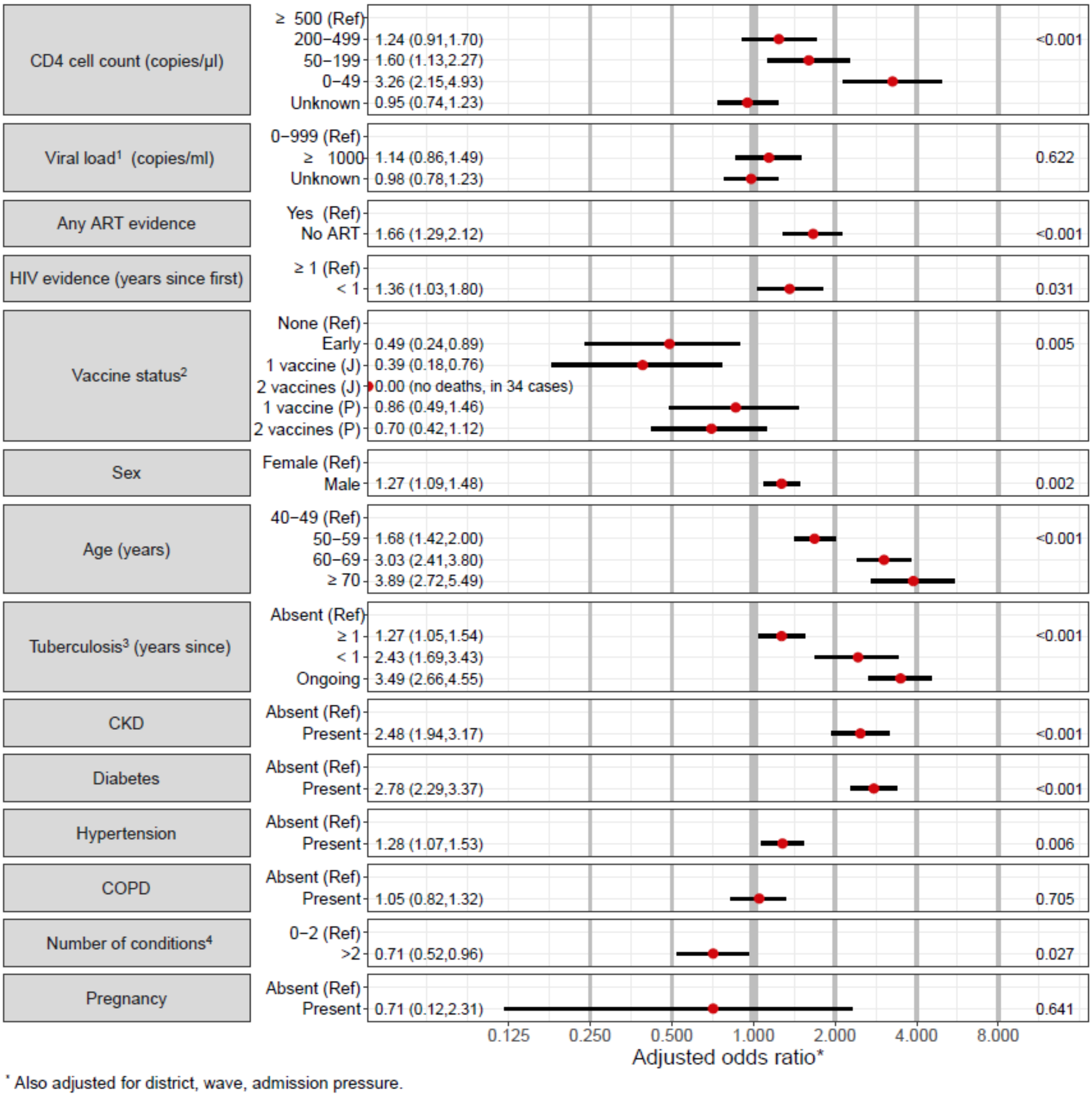


[1] Among those with evidence of ART at any time. [2] The highest level of protection from among six categories: No vaccine received ('None'); <28 days since a first vaccine dose ('Early'); ≥28 days since a first dose of a vaccine, by type ('1 vaccine'); or ≥14 days since a second dose, by type ('2 vaccines'). J: Janssen, P: Pfizer. [3] Time since the most recent evidence of a start of an episode; with episodes beginning after 2 months before the SARS-CoV-2 diagnosis considered as ongoing. [4] Hypertension, diabetes, chronic obstructive pulmonary disease, chronic kidney disease, tuberculosis (at any time).
ART: Antiretroviral therapy. CI: Confidence interval. CKD: Chronic kidney disease. COPD: Chronic obstructive pulmonary disease.
